# Spared Motor Neurons Enable the Control of a Robotic Sixth-Finger for Assistive Grasping in Tetraplegia

**DOI:** 10.1101/2025.02.07.25321673

**Authors:** Daniela Souza de Oliveira, Dominik I. Braun, Maria Pozzi, Leonardo Franco, Matthias Ponfick, Monica Malvezzi, Gionata Salvietti, Domenico Prattichizzo, Alessandro Del Vecchio

**Affiliations:** Department Artificial Intelligence in Biomedical Engineering, Friedrich-Alexander-Universität Erlangen-Nürnberg; Erlangen, Germany; Department of Information Engineering and Mathematics, Università degli Studi di Siena; Siena, Italy; Querschnittzentrum Rummelsberg, Krankenhaus Rummelsberg GmbH; Schwarzenbruck, Germany

## Abstract

Recovering hand function is among the highest priorities for individuals with tetraplegia. Yet, current treatments restoring basic hand movements remain limited for individuals with motor complete spinal cord injury. In this study, we present a non-invasive neuromechatronic interface that directly translates spared motor neuron activities that once encoded the opening and closing of the hand to a supernumerary robotic sixth finger. We re-enabled three individuals with chronic (>8 years) motor complete cervical spinal cord injury to grasp objects important for daily living with the same neural input that controlled the flexion and extension of the fingers. After a few minutes of training, the participants intuitively modulated the discharge activity of their motor units, controlling hand opening and closing. These motor units were then used to control the robotic sixth finger proportionally. All participants successfully executed various grasping tasks that required considerable force from the digits, e.g., opening a bottle by unscrewing the cap. Our findings present a transformative step in assisting hand function, offering an intuitive and non-invasive neuromechatronic interface without the need to learn new motor skills, as the participants use the same motor commands as before the injury. This can significantly improve the quality of life of individuals with paralysis.

## INTRODUCTION

A key focus in restoring hand function is the activity of spinal alpha motor neurons, the last motor pathway of the neuromuscular system. It is known that even individuals with a spinal cord injury (SCI) classified as complete, may retain some spared neural connections above and below the injury level^1–4^. In a previous study involving individuals with motor-complete SCI (eight participants with C5-C6 injury level), we demonstrated that motor units are task-modulated through a non-invasive neural interface using high-density surface electromyography (HDsEMG), enabling the decoding of finger movements^2^. All participants showed distinct associations between specific groups of motor units and specific hand movements so that each movement of the hand digits could be decoded directly. In this previous study, however, the participants controlled a virtual hand with motor unit feedback and were not yet connected to any assistive device.

In this work, we connected the participants spared motor neuron activity to a mechatronic device for the first time. The spared motor neurons that once controlled the natural grasping function of the hand are used here as a signal to control a supernumerary robotic sixth finger to enable basic grasping functions important for daily living^5,6^. Three participants with chronic motor complete SCI, level C5-C6, who also took part in our previous study^2^, were involved. Our neuromechatronic system enabled these individuals to interact again with different objects and perform bimanual tasks like opening bottles or jars, using HDsEMG signals and the robotic sixth finger.

The robotic sixth finger is a supernumerary artificial limb designed to add new degrees of freedom to the human body structure^7,8^, instead of replacing or enhancing human limbs like prostheses and exoskeletons. While for an able-bodied user, a supernumerary robotic finger may improve the ability in complex manipulation tasks like grasping large objects or playing an instrument^9–12^, in the case of people with upper limb disabilities, it has an important application in compensating hand function^5,13,14^. So far, most of the proposed supernumerary robotic limbs are controlled by specific gestures, motions, or muscle activations that must be learned by the user to control the finger (for example, forehead muscles using EMG^5^ and toe control using pressure sensors^12^). Therefore, the main advantage of our proposed system is that there is no need to learn or redirect another motor or muscular function. Specifically, when the participants attempt opening and closing the hand, their real-time motor unit activity provides an intuitive and simple signal indicative of their movement intention, which is used to control the robotic finger. The neural signals the participants use to control the robotic finger are only functionally lost due to their hand paralysis, but these signals can still be read and used to decode their movement intention. This neuromechatronic interface can re-enable hand function for individuals with SCI and opens new possibilities for them to regain control and perform tasks of daily living.

## RESULTS

We developed a non-invasive neuromechatronic system consisting of real-time decoded motor neuron activity and a robotic sixth finger for tetraplegic individuals. We recruited three participants with chronic motor complete SCI (> 8 years after injury). Participants P1 and P2 have bilateral hand paralysis and present good mobility of the arm and wrist. Participant P3 has paralysis in the right hand, with no wrist function and limited arm mobility. Figure 1 shows the concept and overview of the study, in which we recorded spared motor neuron activity using high-density surface electromyography (HDsEMG) as a neural interface and used this activity to control a robotic sixth finger. The neural interface recorded the activity of the extrinsic hand muscles, responsible for hand digit control, using two HDsEMG grids of 64 electrodes each. We decoded this muscle activity by applying a real-time decomposition method that extracted the contributions of individual motor unit action potential spike trains (Fig. 1a, Fig. 2). From these neural ensembles, we obtained a single control signal that was used by the mechatronic device to proportionally control the opening and closing of the sixth finger (Fig. 1b, Fig. 2).

**Fig. 1.**
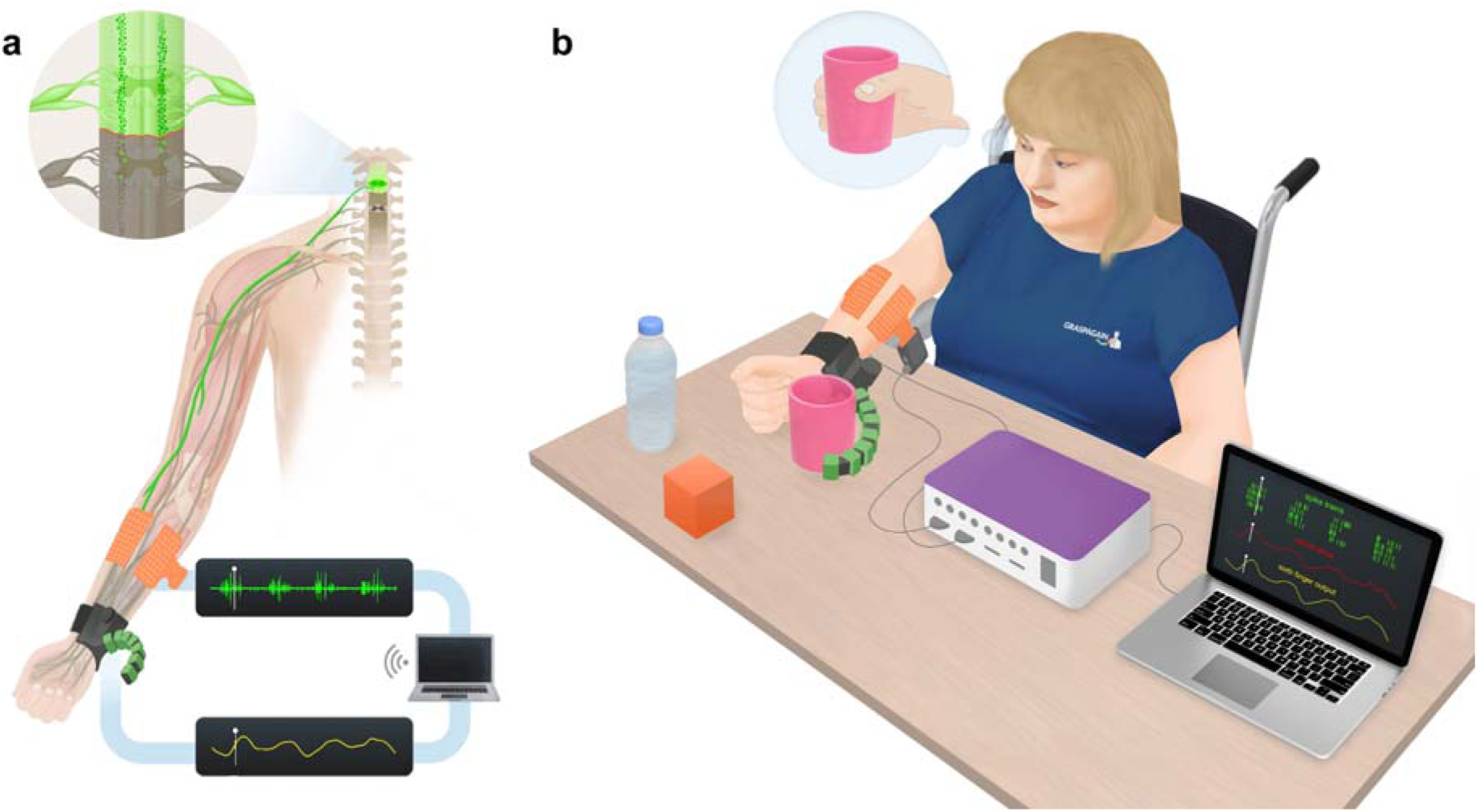
Overview of robotic sixth finger control using spared motor unit activity. (**a**) Representative activation of spared spinal motor neurons to control the robotic sixth finger, using high-density surface electromyography (HDsEMG). (**b**) Participant with cervical SCI with two electrode grids (in orange) placed on the forearm muscles. The high-density surface electromyography signals are recorded while the participant attempts to open and close their hand. These signals are then decomposed into individual motor units, and the cumulative spike train of these motor units is smoothed, resulting in a signal sent as a command to the sixth finger. The sixth finger is then activated according to the motor unit cumulative spike trains, allowing the participant to grasp the objects.

**Fig. 2.**
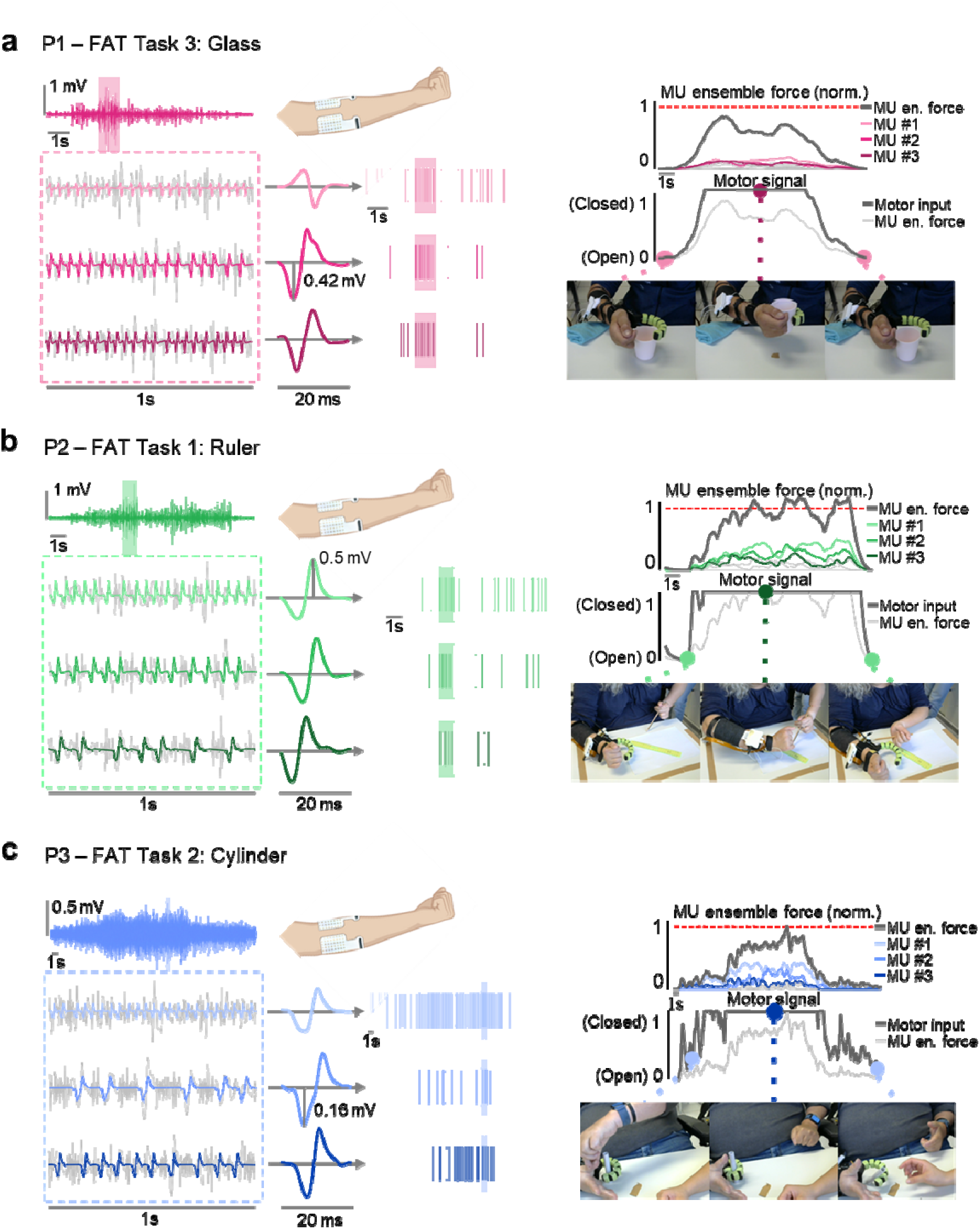
Examples of the neuromechatronic interface in use for each participant. (a) **(b) (c)** Each participant, respectively P1, P2 and P3, performing a different task from the Frenchay arm test (FAT): (**a)** grasping a glass, (**b)** stabilizing a ruler, (**c)** grasping a cylinder. The raw HDsEMG signals from the forearm muscles are first decomposed offline. The motor units identified offline can be detected during the online decomposition step, which allows the control of motor unit activity in real-time. The cumulative spike trains of these motor units identified in real-time are smoothed and this signal is then sent via Wi-Fi and Bluetooth to control the opening and closing of the robotic sixth finger.

To assess the feasibility and assistive relevance of control of the sixth finger by the participants, we asked them to perform various tasks with and without the sixth finger and evaluated their performance. The tasks were divided into three groups: Frenchay arm test^15,16^, pick-and-place tasks (grasping a bottle and using it to pour water, grasping a glass and drinking from it, Box and Blocks test), and bimanual tasks (stacking and unstacking glasses and blocks, opening a spray can and a spread jar). Figure 3 shows the results obtained by the participants with and without the sixth finger, whereas Figure 4 shows some of the tasks included in the protocol. The participants performed the tasks without the sixth finger only once, while tasks executed with the help of the sixth finger required some initial training and once they succeeded in the task, the tasks were repeated 1-2 times. More details on the experimental protocol can be found in the Methods section and Supplementary Figure 1.

**Fig. 3.**
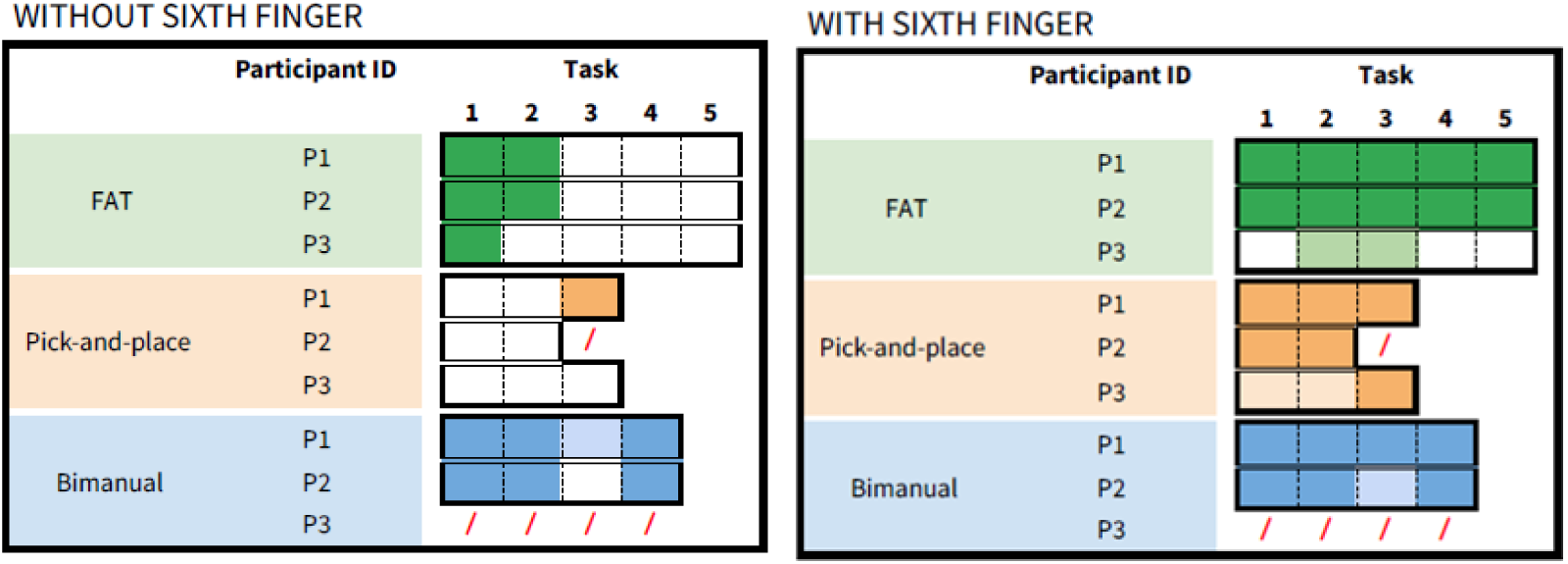
Performance of the three participants in the proposed tasks with and without the robotic sixth finger. Blank spaces indicate failure to complete the task. Red bars represent tasks that were not executed due to participants’ conditions. Lighter colors indicate partial success in the task (See bimanual task number 4 of participant P2 when using the sixth finger).

**Fig. 4.**
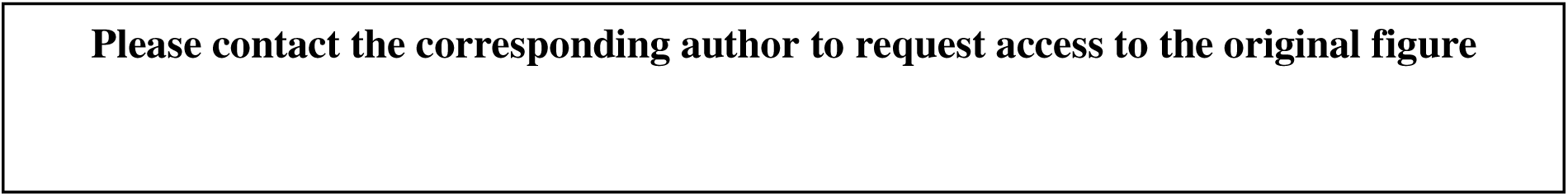
Examples of successful pick-and-place tasks performed by participant P1. (**a)** Task 1: Pouring water into a glass. (**b)** Task 2: Drinking from a glass and placing the glass over the table upside-down.

We found that all three participants could control the sixth finger using the same neural inputs that once controlled hand opening and closing, allowing them to execute most of the proposed tasks. As anticipated, there was a clear difference in performance according to the arm and wrist mobility of the participants, so participants who showed better arm mobility could complete the tasks more efficiently. Videos of participants P1 and P2 executing the tasks are shown, respectively, in Supplementary Videos 1 and 2.

As shown in Figure 3, both participants, P1 and P2, achieved similar scores in most tests. For the Frenchay arm test, they were able to stabilize a ruler and grasp a cylinder (tasks 1-2) with the sixth finger and without it, using one hand. The tasks 3-5, picking up a glass, removing and replacing a sprung clothes peg, and combing hair were successfully performed by participants P1 and P2 with the sixth finger, while without the sixth finger, these tasks could only be performed using both hands. The same was observed for the pick-and-place tasks, which were mostly re-enabled due to the sixth finger.

For participant P3, in the Frenchay arm test, the participant could stabilize the ruler without the robotic finger but not with it (this was due to a problem with the height of the sixth finger when touching the table). For the pick-and-place tasks, the participant could hold a bottle and grasp a glass, but due to the low mobility of the arm, he was not able to lift the arm to ‘drink’ or turn the arm to put the glass upside down on the table.

Regarding the Box and Block Test, a standard assessment of unilateral manual dexterity ^17^, participant P1 was able to perform the test both without and with the sixth finger, while P3 could only perform it with the sixth finger. Participants were asked to transfer as many small wooden blocks as possible from one side of the working space to the other in one minute. Participant P1 could transfer 5 blocks/minute with the sixth finger and 14 blocks/minute without it. P1 is a well-adapted participant, he has passive hand function and developed compensatory strategies for performing hand tasks. Therefore, the difference in task execution time was expected as he was using the sixth finger for the first time and experienced some delays due to latencies of the system and had some difficulty in relaxing the muscles to re-open the robotic finger. Participant P3 was also able to complete the box and blocks test, transferring 4 blocks/minute.

Additionally, four bimanual tasks were presented to participants, including stacking/unstacking glasses, removing the lid from a jar, removing the lid from a spray can, and stacking/unstacking Lego DUPLO blocks. Bimanual tests were not performed for participant P3 since his left hand is fully functional.

For participants P1 and P2, the only bimanual task they were unable to complete without the sixth finger was opening the spray can, which required more force than the other tasks. P1 showed he could still complete this task without the sixth finger, by using both hands and pressing the can against his body to open it. P2 could complete this task using the sixth finger with assistance (another person held the can and P2 opened it), due to a lack of strength in her left hand, but she also did not attempt the task outside of the delimited working space.

After the tests were completed, the participants answered a system’s usability scale (SUS) survey, the results were 52.5 for P1, 60 for P2, and 65 for P3. Figure 5 presents the participants’ answers to each question. They found the neuromechatronic interface to be not complex, well integrated, and quick to learn, requiring minimal learning to use it. Conversely, all participants noted they would need the support of a technical person, found the system cumbersome to use, and reported some inconsistencies. Considering the actual version of our system is not wireless, with an EMG amplifier, cables, and grids of electrodes, these results were expected, since this limited some of the movements.

**Fig. 5.**
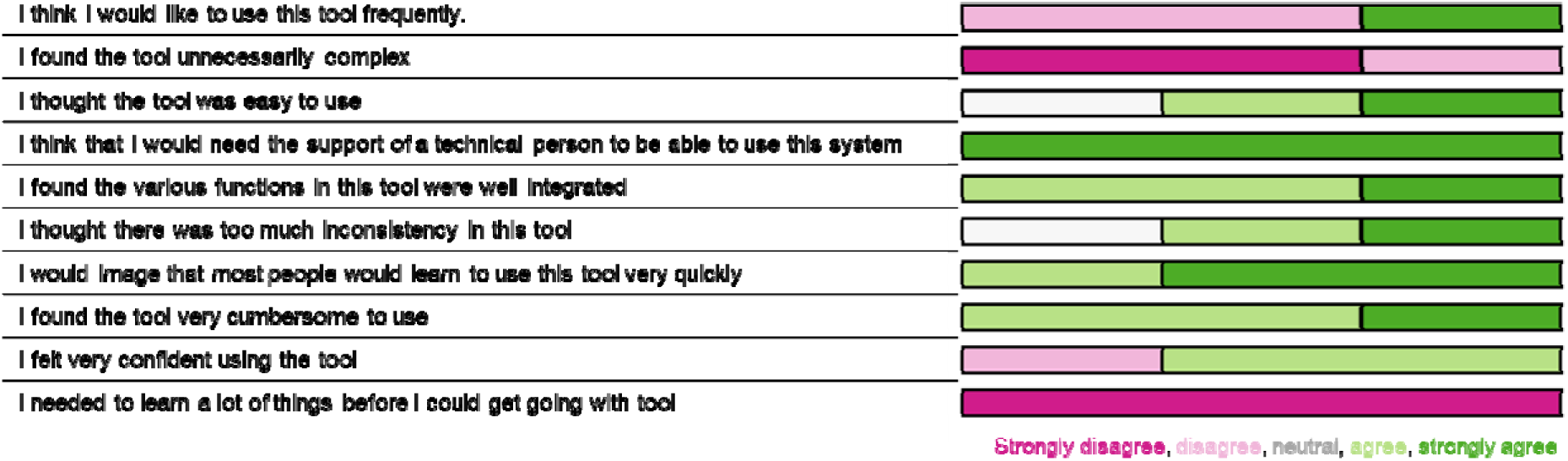
Evaluation of the interface. System usability scale (SUS) feedback from the three participants on the neuromechatronic interface.

## DISCUSSION

Our findings demonstrated that the participants could effectively control and use the robotic sixth finger to manipulate and grasp objects of daily use. This accomplishment is particularly remarkable given that these are individuals with chronic motor complete SCI and that they were proportionally controlling the robotic finger with their spared neural activity, encoding the opening and closing of the hand. The sixth finger, integrated with the neural interface, emerged as a promising tool to re-enable basic grasping function.

There were some restrictions on the tasks the participants could perform, depending on their arm and wrist mobility. For participants, P1 and P2, with both hands paralyzed, limited to only passive function, but with good arm mobility, the advantage of using the sixth finger is to allow them to perform tasks with only one hand and with better dexterity. For participant P3, with much less mobility of the right arm, only a few tasks could be completed, but those were only possible by using the sixth finger. Overall, the functional advantage of the sixth finger was evident in tasks requiring enough grip strength to be able to lift objects without dropping them.

Comparing the performance with and without the sixth finger is difficult, since these participants have been dealing with the injury for years and were trained to execute tasks by using a passive grasp, while here they used the sixth finger mostly for the first time. Given that, we are also aware that our neuromechatronic system presents some limitations, especially when considering the survey results. The reported inconsistencies are related to the responsiveness of the system (due to the latency in the connection between the neural interface and the sixth finger) and the low velocity of the extension motion of the sixth finger. This last issue might have been influenced by the participants’ self-reported difficulty in relaxing their muscles, likely due to involuntary muscle contractions^18^ and/or the possible problems with the online decomposition. Another issue is that the current version of our system is not fully wearable and wireless, which might have constrained some of the movements. Those issues will be addressed in the future to improve our system. Moreover, when answering open-ended questions after the SUS, the participants expressed interest in a wearable version of the system and reported that they did not have to learn new skills to use it, finding it simple to control, which confirms the system is intuitive and no additional motor command is necessary to use the sixth finger.

The use of HDsEMG is essential due to its spatial resolution, which allows for more accurate detection of spared motor neuron activity, leading to improved control. In fact, even signals from a single motor unit can be sufficient for this control. In the future, HDsEMG electrodes could be used to control additional degrees of freedom or to selectively place bipolar electrodes to reduce the need for more electrodes and provide a simple wearable version.

It is important to note that the intention of using the robotic finger is not to replace a limb, but to offer a simple solution to re-enable grasping function. This differs from using an exoskeleton, for example. A challenge in designing exoskeletons for individuals with SCI is dealing with a tenodesis hand^19^, where the tendons are surgically shortened to allow a passive grasp. At the same time, this limits the range of motion of the finger when using an exoskeleton^20^. Restoring the length of the tendon is not trivial and requires surgery. Thus, the sixth finger represents a way to bypass muscle and tendon limitations to re-enable grasping. Additionally, the sixth finger could be useful as a rehabilitation tool, enabling individuals with SCI to practice grasping and, indirectly, movements of the arm, as well as practice individual finger movements with visual feedback from it. In summary, by combining HDsEMG, with real-time motor unit decomposition, and a supernumerary robotic sixth finger, we demonstrated a natural and intuitive control of the robotic finger by participants with chronic motor complete SCI.

## MATERIALS AND METHODS

### Participants

For this study, we recruited three participants with chronic motor complete SCI, with injury level C5-C6. Participants’ characteristics are provided in Table 1. Participants have different arm and wrist mobility, with P1 and P2 being able to lift their arm and move the wrist, and P3 only paralyzed in the right hand, with limited arm mobility and unable to move the wrist. All the procedures were performed according to the Declaration of Helsinki and approved by the Friedrich-Alexander Universität ethics committee (Number 24-208-B). Participants signed a written informed consent.

**Table 1.**
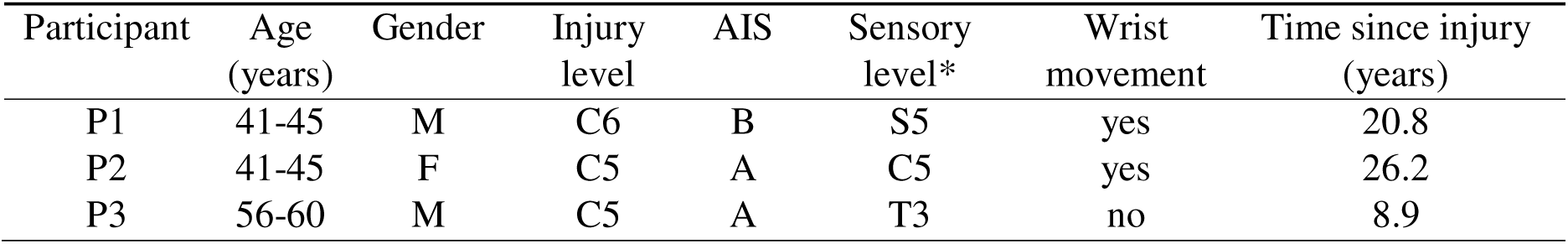
Characteristics of recruited participants in the study. *The sensory level corresponds to the lowest level with normal sensory function.

### Overview of the study and protocol

This study was conducted in one experimental session that was 2-3 hours long. First, the participants were familiarized with the neural and mechatronic interface. As a warm-up, we asked the participants to follow the movements of a virtual hand opening and closing the fingers on a video in front of them. The duration of the video was 42 s, and it was played twice. Subsequently, we asked the participants to attempt the same movement task again and concurrently recorded 20 s of HDsEMG signals from their forearm muscles for our offline motor unit decomposition. We then tested if the participants could control their motor unit activity in real-time with an online decomposition.

To test the real-time control of the robotic finger integrated with the neural interface, we first placed the robotic finger on a table in front of the participants. We asked them to attempt opening and closing their fingers and to observe if they were also controlling their motor unit activity (visual feedback on a screen) and the robotic finger accordingly. Once these procedures were completed and fully tested and the participants were comfortable with the neuromechatronic interface, we strapped the robotic finger to the participants’ wrists. A short training phase followed, in which the participants practiced grasping some objects, such as a glass, or a cube.

We then began a series of tests to assess if the participants would be able to perform tasks of daily living by controlling the robotic finger. We separated our protocol into three groups of tasks: Frenchay arm test, pick-and-place, and bimanual tasks described below. We assessed the feasibility of the tasks with and without the neuromechatronic interface by evaluating their performance (failure or successful completion of a task). The tasks without the robotic finger were performed only once. Due to the non-familiarity with the protocol and the robotic finger, we allowed the participants a short training time to attempt the tasks with the sixth finger, and only after that, we evaluated if they succeeded or not on the task. At the end of the experimental session, we asked participants to answer a survey using the standardized system usability scale (SUS). The tasks of the protocol are depicted in Supplementary Figure 1.

### Frenchay arm test

We first wanted to verify the participants’ performance with a standardized test. The Frenchay arm test is a well-known test^21^, consisting of five tasks performed with the dominant arm.

Task 1: Stabilize a ruler and draw a line with a pencil in the other hand. The ruler must be held firmly.

Task 2: Grasp a cylinder (12 mm diameter, 5 cm long) placed 15 cm from the edge of the table, lift it, and return it in place without dropping it.

Task 3: Pick up a glass, positioned 15 cm from the edge of the table, drink and return it in place.

Task 4: Remove and replace a sprung clothes peg from a 10 mm diameter dowel, 15 cm long set in a 10 cm base, 15 cm from the edge of the table.

Task 5: Comb hair imitation, across the top of the head.

### Pick-and-place tasks

Task 1: Grasp a bottle, pour water into a glass, and return the bottle in place.

Task 2: Grasp a glass, drink, and return it in place upside down.

Task 3: Box and block test – moving blocks from one side of the table/workspace to the other. We recorded how many blocks were transferred in one minute.

### Bimanual tasks

These tests required the use of both hands, with the robotic finger on the right hand, the participants could then use their left hand to help.

Task 1: Unstack and stack glass.

Task 2: Remove the lid of a spread jar.

Task 3: Remove the lid from a spray can.

Task 4: Stack and unstack DUPLO blocks.

### Survey

After the tests, the participants answered the questions from the system usability scale (SUS)^22^ and two additional questions related to their perception of the neuromechatronic interface: “Would you use a wearable version of the setup at home?” and “Do you have any suggestions on how to improve the system?”.

### Evaluation of task performance

To compare the tasks with and without the robotic finger, for the Frenchay arm test, we considered a successful task, if, after familiarization, the task could be performed once. For pick-and-place and bimanual tasks, we considered a successful task, if the task could be performed at least twice (1x training, 1x repetition). For the box and blocks, we compared how many blocks per minute were successfully transferred across the table. Regarding the survey, SUS results were converted to the appropriate scale and a score was calculated for each participant. We also presented the results for each question (Fig. 5).

### HDsEMG recordings

We placed two grids of electrodes (8×8, interelectrode distance = 10 mm) on the forearm muscles of the participants, aligned to the ulna bone, covering the flexor and extensor muscles. The skin of the forearm was shaved and cleansed with 70% ethyl alcohol. We attached the electrode grids to the skin using bi-adhesive foams and conductive paste (SpesMedica, Battipaglia, Italy). Two reference electrodes were placed, one on the elbow joint and the other on the styloid process of the ulna. We recorded and streamed the HDsEMG signals in monopolar mode with a multichannel amplifier 16-bit A/D converter (Quattrocento, OT Bioelettronica), sampling frequency of 2048 Hz, and bandpass filter of 10-500 Hz. For the streaming of the HDsEMG signals, the protocol TCP/IP was used, and the signals were sent to our decomposition software through the software OT BioLab Light (OT Bioelettronica) with a refresh frequency of 32 Hz.

### Neural interface

The neural interface used in the study, designed for real-time control of a neuromechatronic system, is based on HDsEMG and motor unit activity. The process consists of two main steps: offline and online decomposition.

During the offline decomposition, HDsEMG signals are recorded from the forearm muscles of participants while they perform a movement task (opening and closing of their digits) as instructed by a virtual hand video on a monitor. These signals are then analyzed using a blind source separation (BSS) technique that combines fast independent component analysis (FastICA) with convolutive sphering. This approach allows for the decomposition of the recorded signals into individual motor units that are involved in the attempted movement. The separation matrix derived from this decomposition is saved for the online decomposition in the next step.

In the online decomposition step, i.e. real-time phase, the predetermined separation matrix is applied to continuously decompose motor unit discharges while participants attempt to control the robotic finger. The decomposed motor unit discharges are converted into spike trains, which are then summed to form a cumulative motor unit spike train. We applied smoothing to the cumulative spike train to generate an input signal for the robotic finger. This is done by convolving the spike train with a physiologically driven motor unit twitch model (total length of 1.5 s)^23^, resulting in a signal referred to as motor unit ensemble force in this paper. Finally, the signal is normalized using the maximum discharge rate obtained during the offline decomposition, allowing for precise and proportional controlled movement of the robotic finger. The details of both steps are described below.

### Offline decomposition

Offline decomposition is a critical step in our real-time application, as it allows us to identify appropriate motor unit filters that are needed to extract the spike trains of individual motor units from HDsEMG signals. These signals can be modeled as the sum of motor unit action potentials and noise. Mathematically, this can be expressed for 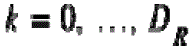 where 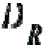is the duration of the recording:

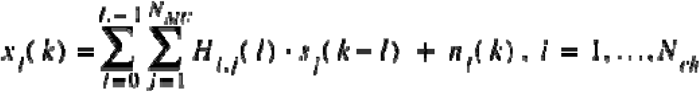

In this expression, 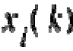 denotes the measured EMG signal in the i-th channel of the electrode grid, with 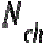 representing the number of electrode channels. The term 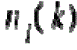 accounts for the additive noise in the respective electrode channel. The spike train 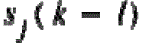 comprises the discharges from j-th motor unit with 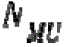 indicating the number of motor units that can be observed within the electrode grid. The motor unit action potential matrix 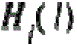 provides information on the action potential waveform for the i-th channel (row) and the j-th motor unit (column) over a length of L.

During the offline decomposition, the goal is to extract information about the term:

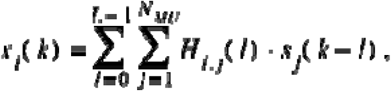

which is unknown during the recording of the raw HDsEMG signals. The individual sources of the HDsEMG, i.e., the motor unit action potentials and their convolution with the corresponding spike trains, are masked by the summation of hundreds of motor unit discharges. To address this, a BSS technique is applied, a method successfully used with HDsEMG in recent decades ^24–28^. For our study, we adapted the convolutive BSS technique proposed by Negro et al., 2016 which showed a highly accurate extraction of motor units in HDsEMG. Details of the applied processing steps are provided below.

After recording, the HDsEMG signals were bandpass filtered between 20-500 Hz using a second-order Butterworth filter, and a 50 Hz notch filter was applied to reduce power line interference. This filtering removes noisy frequencies that do not carry significant information. Afterwards, the filtered HDsEMG signals are extended to increase the number of observations for the BSS technique by adding *R-*delayed versions of each electrode channel. Here, *R* = 4 is calculated using 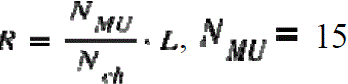 components, 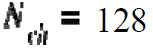 electrode channels, and L = 40 samples (estimated motor unit action potential duration of 0.02 s with a sampling frequency of 2048 Hz). After extension, the data is centered by subtracting the mean within each electrode channel to ensure zero mean data, which is crucial for accurate processing. Finally, the extended and centered observations are whitened. Whitening transforms the data such that the covariance matrix of the extended observations equals the identity matrix at time lag zero. This procedure decorrelates the data and standardizes the variance, reducing dependencies between signals and thus lowering the computational complexity of the source separation process.

After convolutive sphering, we applied a fixed-point algorithm known as FastICA, which includes a Gram-Schmidt orthogonalization step. In our approach, FastICA operates iteratively using a deflation method, meaning that the sources are identified one by one. During the offline step, we aimed to identify 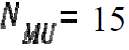 motor units, resulting in 15 distinct innervation pulse trains (IPTs), i.e., the source signals.

In addition to the source signals, the separation matrix was also obtained and saved for the online decomposition. This separation matrix is crucial because it can be used to extract the IPTs from the HDsEMG during online operation, bypassing the need for FastICA, which is computationally intensive and unsuitable for real-time processing.

Once the IPTs had been identified, a spike detection process was applied to decode the motor unit spike trains. Subsequently, a K-means clustering approach, driven by silhouette scores, was employed to categorize the peaks of the IPTs into two distinct groups: higher peaks and lower peaks. Peaks of a lower amplitude were considered unimportant and thus discarded. Conversely, peaks of a higher amplitude with a silhouette score of 0.9 or above were considered to be valid motor unit discharges.

As a final step in the offline decomposition, we calculate the motor unit ensemble force from the cumulative spike train of the decomposed motor units. This is done by convolving the cumulative spike train with a physiologically driven motor unit twitch model ^23^. This model simulates a muscle twitch and includes a latent period, a contraction phase, and a relaxation phase. The maximum value of the motor unit ensemble force is then extracted and stored for use in real-time applications.

### Online decomposition (real-time control of motor unit activity)

In contrast to offline decomposition, the online decomposition step faces different challenges due to time constraints and computational load. To ensure smooth operation, the latency between the participant’s motion intent and the execution of movement by the neuromechatronic device should not exceed 100-175ms^29^. The high computational demands of BSS techniques, such as whitening and FastICA, make them unsuitable for real-time processing.

In offline, the entire 20 s recording is decomposed, while in real-time, the system processes 32 frames of HDsEMG data, each frame containing 64 samples per channel. To simplify the online decomposition, we apply only the extension and centering steps of convolutive sphering. The resulting signal is then multiplied by the precomputed separation matrix to obtain the IPTs.

For spike detection in real-time, we identify peaks in the IPTs and apply a threshold based on the baseline noise recorded in the first frame of the real-time protocol, with the participant’s hand at rest. The default threshold is set to 10 times the baseline noise but can be adjusted through the graphical user interface to enhance sensitivity. To improve spike detection reliability, we use template matching to compare the motor unit action potential shapes obtained from the offline step with those detected in real-time. A motor unit is considered valid if its correlation coefficient with the template is 0.6 or higher.

The estimated motor unit ensemble force is calculated similarly to the offline decomposition. Given the relatively long duration of the motor unit twitch (1.5 s), we buffer the motor unit ensemble force and add its new estimation from the current frame to the buffer. The average motor unit ensemble force of the current frame is then used to calculate the input control signal for the robotic sixth finger. Finally, the maximum value of the motor unit ensemble force stored during the offline step is used to normalize the control signal, ensuring it stays between 0 and 1. If the normalized force exceeds 1, it is bound to 1 to prevent the robotic finger from surpassing its motor limits.

### Mechatronic interface

The supernumerary finger used in this paper is a wearable device composed of a robotic finger with a soft-rigid structure and a base that can be strapped to the user’s wrist. The finger is tendon-driven and can be composed of a variable number of modules comprising a rigid part (phalanx), and a soft part (joint). Compared to previous versions of the robotic sixth finger^5^, the adopted device has a higher friction at the phalanges due to silicone rubber covers (shown in green, Fig. 1), and a more rounded design to improve safety and comfort. In addition, the robotic finger is easy to assemble due to a new tendon routing design, where the tendon channel extends all the way through the dorsal side of the module, allowing the easy addition/removal of modules. Also, the flexible joints are made of two identical parts, facilitating their insertion in the rigid parts. The silicone covers have been created by pouring 1:1 Super Rapid Silicone Rubber, R PRO FAST by Reschimica, which is non-toxic and resistant to breakage, in a 3D-printed mold made of Acrylonitrile butadiene styrene (ABS). The rigid phalanges have been built in ABS, whereas the compliant joints are made of Thermoplastic polyurethane (TPU). The robotic finger used in this study has 7 modules that are moved all together through a tendon pulled by a single electric servomotor. The base of the finger supports the motor and has an ergonomic shape so that it can be worn on the user’s wrist through a fabric band with velcro tape. The finger weighs 154 g and is 151 mm long.

### Integration of neural and mechatronic interfaces

We have implemented a User Datagram Protocol (UDP) network protocol to facilitate communication between the neural and mechatronic interfaces, enabling intuitive control to the user. LabVIEW software (Version 18.0, National Instruments, Austin, TX, USA) was used to bridge the UPD incoming data stream to the microcontroller of the supernumerary finger via Bluetooth. To minimize the risk of buffer overflow and prevent overstraining the actuation system, we streamed only every third frame of the normalized motor unit ensemble force to the mechatronic device. This approach resulted in a streaming frequency of approximately 10.66 Hz.

## Supplementary Materials

Supplementary figures: Fig. S1

Supplementary movies: Movies S1 to S3

## Data Availability

All data produced in the present study are available upon reasonable request to the authors.

## Acknowledgments

We thank the participants who contributed to this study.

## Funding

European Research Council (ERC) Starting Grant project GRASPAGAIN 101118089 (ADV)

German Ministry for Education and Research (BMBF) through the project MYOREHAB under Grant 01DN2300 (ADV)

Bavarian State Ministry of Economic Affairs and Media, Energy and Technology through the project NeurOne under grant LSM-2303-0003 (ADV)

Bavarian State Ministry of Economic Affairs and Media, Energy and Technology through the project GraspAgain under grant MV-2303-0006 (ADV)

Dhip (Digital Health Innovation Platform) campus-bavarian aim (ADV and DSO) European Union by the Next Generation EU project ECS17 “THE – Tuscany Health Ecosystem PNRR MUR M4 C2 Inv. 1.5, CUP B63C22000680007, Spoke 9 – (MP, LF, GS and DP)

Horizon Europe project “HARIA – Human-Robot Sensorimotor Augmentation – Wearable Sensorimotor Interfaces and Supernumerary Robotic Limbs for Humans with Upper-limb Disabilities” under grant 101070292 (MP, LF, GS and DP)

## Data and materials availability

All data, code, and materials used in the present study are available upon request to the authors. An executable version of the decomposition software is accessible here: github.com/NsquaredLab/NeurOne.

## References and Notes

1. Ting, J. E., et al. Sensing and decoding the neural drive to paralyzed muscles during attempted movements of a person with tetraplegia using a sleeve array. J. Neurophysiol. 126, 2104– 2118 (2021).

2. Souza Oliveira, D., et al. A direct spinal cord–computer interface enables the control of the paralysed hand in spinal cord injury. Brain awae088 (2024) doi:10.1093/brain/awae088.

3. Sharma, P., et al. Preservation of functional descending input to paralyzed upper extremity muscles in motor complete cervical spinal cord injury. Clin. Neurophysiol. 150, 56–68 (2023).

4. Sherwood, A. M., Dimitrijevic, M. R. & Barry McKay, W. Evidence of subclinical brain influence in clinically complete spinal cord injury: discomplete SCI. J. Neurol. Sci. 110, 90– 98 (1992).

5. Hussain, I., Salvietti, G., Spagnoletti, G. & Prattichizzo, D. The Soft-SixthFinger: a Wearable EMG Controlled Robotic Extra-Finger for Grasp Compensation in Chronic Stroke Patients. IEEE Robot. Autom. Lett. 1, 1000–1006 (2016).

6. Salvietti, G., et al. Compensating Hand Function in Chronic Stroke Patients Through the Robotic Sixth Finger. IEEE Trans. Neural Syst. Rehabil. Eng. 25, 142–150 (2017).

7. Prattichizzo, D., et al. Human augmentation by wearable supernumerary robotic limbs: review and perspectives. *Prog*. Biomed. Eng. 3, 042005 (2021).

8. Dominijanni, G., et al. The neural resource allocation problem when enhancing human bodies with extra robotic limbs. *Nat*. Mach. Intell. 3, 850–860 (2021).

9. Parietti, F. & Asada, H. Supernumerary Robotic Limbs for Human Body Support. IEEE Trans. Robot. 32, 301–311 (2016).

10. Sasaki, T., Saraiji, M. Y., Fernando, C. L., Minamizawa, K. & Inami, M. MetaLimbs: multiple arms interaction metamorphism. in *ACM SIGGRAPH 2017 Emerging Technologies* 1–2 (ACM, Los Angeles California, 2017). doi:10.1145/3084822.3084837.

11. Shafti, A., Haar, S., Mio, R., Guilleminot, P. & Faisal, A. A. Playing the piano with a robotic third thumb: assessing constraints of human augmentation. Sci. Rep. 11, 21375 (2021).

12. Kieliba, P., Clode, D., Maimon-Mor, R. O. & Makin, T. R. Robotic hand augmentation drives changes in neural body representation. *Sci*. Robot. 6, eabd7935 (2021).

13. Ort, T., Wu, F., Hensel, N. C. & Asada, H. H. Supernumerary Robotic Fingers as a Therapeutic Device for Hemiparetic Patients. in *Volume 2: Diagnostics and Detection; Drilling; Dynamics and Control of Wind Energy Systems; Energy Harvesting; Estimation and Identification; Flexible and Smart Structure Control; Fuels Cells/Energy Storage; Human Robot Interaction; HVAC Building Energy Management; Industrial Applications; Intelligent Transportation Systems; Manufacturing; Mechatronics; Modelling and Validation; Motion and Vibration Control Applications* V002T27A010 (American Society of Mechanical Engineers, Columbus, Ohio, USA, 2015). doi:10.1115/DSCC2015-9945.

14. Hussain, I., Spagnoletti, G., Salvietti, G. & Prattichizzo, D. Toward wearable supernumerary robotic fingers to compensate missing grasping abilities in hemiparetic upper limb. Int. J. Robot. Res. 36, 1414–1436 (2017).

15. 15. Katie Marvin. Frenchay Arm Test (FAT). https://strokengine.ca/en/assessments/frenchay-arm-test-fat/ (2012).

16. Wade, D. T., Langton-Hewer, R., Wood, V. A., Skilbeck, C. E. & Ismail, H. M. The hemiplegic arm after stroke: measurement and recovery. J. Neurol. Neurosurg. Psychiatry 46, 521–524 (1983).

17. Mathiowetz, V., Volland, G., Kashman, N. & Weber, K. Adult Norms for the Box and Block Test of Manual Dexterity. Am. J. Occup. Ther. 39, 386–391 (1985).

18. Zijdewind, I. & Thomas, C. K. Firing patterns of spontaneously active motor units in spinal cord injured subjects. J. Physiol. 590, 1683–1697 (2012).

19. Fridén, J., House, J., Keith, M., Schibli, S. & Van Zyl, N. Improving hand function after spinal cord injury. J. Hand Surg. Eur*. Vol.* 47, 105–116 (2022).

20. Bagneschi, T., et al. A Soft Hand Exoskeleton With a Novel Tendon Layout to Improve Stable Wearing in Grasping Assistance. IEEE Trans. Haptics 16, 311–321 (2023).

21. Heller, A., et al. Arm function after stroke: measurement and recovery over the first three months. J. Neurol. Neurosurg. Psychiatry 50, 714–719 (1987).

22. SUS: A ‘Quick and Dirty’ Usability Scale. in (eds. Jordan, P. W., Thomas, B., McClelland, I. L., Weerdmeester, B. & Brooke, John) 207–212 (CRC Press, 1996). doi:10.1201/9781498710411-35.

23. Braun, D. I. et al. NeurOne: High-performance Motor Unit-Computer Interface for the Paralyzed. Preprint at 10.1101/2023.09.25.23295902 (2023).

24. Holobar, A. & Farina, D. Blind source identification from the multichannel surface electromyogram. Physiol. Meas. 35, R143–R165 (2014).

25. Holobar, A. & Farina, D. Noninvasive Neural Interfacing With Wearable Muscle Sensors: Combining Convolutive Blind Source Separation Methods and Deep Learning Techniques for Neural Decoding. IEEE Signal Process. Mag. 38, 103–118 (2021).

26. Holobar, A. & Zazula, D. Gradient Convolution Kernel Compensation Applied to Surface Electromyograms. in Independent Component Analysis and Signal Separation (eds. Davies, M. E., James, C. J., Abdallah, S. A. & Plumbley, M. D.) vol. 4666 617–624 (Springer Berlin Heidelberg, Berlin, Heidelberg, 2007).

27. Holobar, A. & Zazula, D. Multichannel Blind Source Separation Using Convolution Kernel Compensation. IEEE Trans. Signal Process. 55, 4487–4496 (2007).

28. Negro, F., Muceli, S., Castronovo, A. M., Holobar, A. & Farina, D. Multi-channel intramuscular and surface EMG decomposition by convolutive blind source separation. J. Neural Eng. 13, 026027 (2016).

29. Farrell, T. R. & Weir, R. F. The Optimal Controller Delay for Myoelectric Prostheses. IEEE Trans. Neural Syst. Rehabil. Eng. 15, 111–118 (2007).

