## Supplementary materials for "Spared Motor Neurons Enable the Control of a Robotic Sixth-Finger for Assistive Grasping in Tetraplegia"

### Supplementary Figures

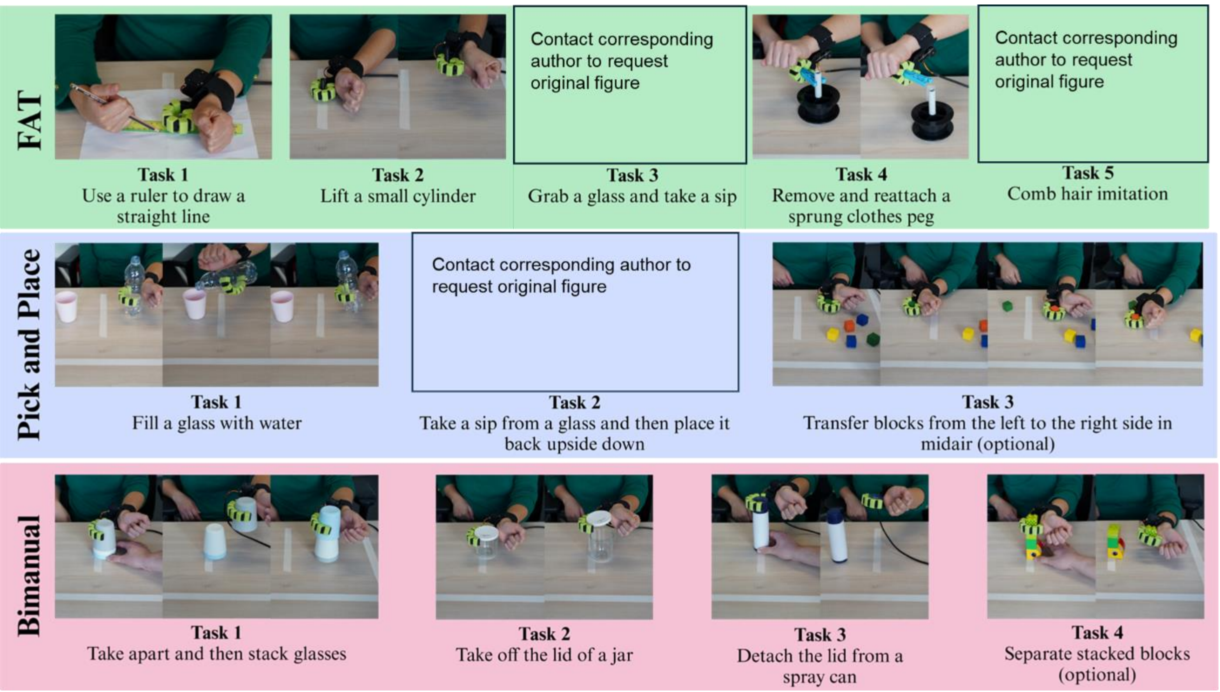


**Supplementary Fig. S1**. Description of protocol tasks

### Supplementary Movies

**Supplementary Movie S1**. Tasks performed by Participant 1 (P1). Includes a selection of videos performed by P1 with and without the robotic sixth-finger.
https://youtu.b/kGu-ChIqakI

**Supplementary Movie S2**. Tasks performed by Participant 2 (P2). Includes a selection of videos performed by P2 with and without the robotic sixth-finger
https://youtu.be/tSEo9s4E43Q

**Supplementary Movie S3.** Summary of the paper. Includes a summary of the paper contribution and shows a few selected videos and results.
https://youtu.be/bDiF3vVyd_4
